# Outcomes of Women with Primary and Second Primary Ovarian Cancer in the United States

**DOI:** 10.1101/2025.06.25.25330320

**Authors:** Oluwasegun Akinyemi, Phiwinhlanhla Ndebele-Ngwenya, Mojisola Fasokun, Terhas Weldeslase, Seun Ikugbayigbe, Eunice Odusanya, Oluebubechukwu Eze, Miriam Michael, Edward Cornwell, Kakra Hughes, Guoyang Luo

## Abstract

**Background:** Ovarian cancer is a leading cause of gynecologic cancer-related mortality. While most cases arise as a primary malignancy, a subset occurs as a second primary cancer following a previous non-ovarian malignancy. Limited studies have examined the survival differences between these groups.

**Objective:** To compare the characteristics, treatment patterns, and survival outcomes of women with primary ovarian cancer versus second primary ovarian cancer and identify factors associated with cancer-specific survival (CSS) and overall survival (OS).

**Methods:** This retrospective cohort study utilized data from the Surveillance, Epidemiology, and End Results (SEER) 18 Registry (2000–2021). Women aged ≥18 years diagnosed with primary or second primary ovarian cancer were included. Multivariable Cox proportional hazards models estimated the association between primary cancer status and survival outcomes, adjusting for demographic, tumor, and treatment-related factors.

**Results:** A total of 27,308 women were included: 23,132 (84.7%) with primary ovarian cancer and 4,176 (15.3%) with second primary ovarian cancer. Women with second primary ovarian malignancies were older (44.6% vs. 35.6% >64 years, p<0.001) and more likely to be White (71.3% vs. 66.0%, p<0.001). They were diagnosed more frequently at earlier stages (Stage I: 35.4% vs. 32.4%, p<0.001). Women with second primary ovarian cancer had significantly better CSS at 5 years (60.3% vs. 56.8%, p<0.001) and 20 years (43.5% vs. 39.1%, p<0.001). After adjustment, they had a 7.2% lower risk of cancer-specific mortality (HR: 0.928, 95% CI: 0.876-0.982, p=0.010). However, OS was similar at 5 years (53.0% vs. 53.5%, p<0.001), with a survival disadvantage emerging over time (HR: 1.057, 95% CI: 1.005-1.113, p=0.031).

**Conclusion:** Women with second primary ovarian cancer had better CSS than those with primary ovarian cancer, likely due to earlier detection and increased surveillance. However, long-term OS disparities suggest a need for continued optimization of follow-up care and management strategies.

## Introduction

Ovarian cancer, often referred to as the silent killer due to its subtle symptoms and late-stage diagnosis[1], remains one of the most lethal gynecologic malignancies[2, 3]. Despite its relatively low incidence[4, 5], ovarian cancer ranks fifth in cancer-related deaths among women[1]. According to the National Cancer Institute’s Surveillance, Epidemiology, and End Results (SEER) Program, ovarian cancer is the eighteenth most common cancer in the United States, with an estimated 19,680 new cases and 12,740 deaths anticipated in 2024[5, 6]. The prognosis for ovarian cancer is heavily stage-dependent, with localized cases exhibiting the highest 5-year survival rates[7], while distant metastases are associated with poor outcomes[5, 8]. Alarmingly, 55% of ovarian cancer cases are diagnosed at an advanced stage, whereas only 19% are detected early when still localized[4, 8].

The mean age at diagnosis is 63 years[9], with the highest incidence rates observed among non-Hispanic American Indian/Alaskan Native and non-Hispanic White women[9, 10]. While hereditary factors play a significant role in ovarian cancer risk[11], the majority of cases arise from unknown etiologies[10]. Breast cancer gene(BRCA) gene mutations significantly increase the likelihood of developing ovarian, breast, fallopian tube, and peritoneal cancers[12]. In addition, hereditary non-polyposis colorectal cancer (HNPCC), caused by mutations in DNA mismatch repair genes, increases the lifetime risk of ovarian cancer by 7-12%[13]. Other tumor suppressor genes, such as TP53 and PTEN, have also been implicated in ovarian cancer development[14]. Beyond genetic predisposition, non-hereditary risk factors include early menarche, infertility, increased age, and nulliparity[15].

Ovarian cancer is a heterogeneous disease with multiple histologic subtypes, which can be classified based on cell origin and site of development[16–18]. The most common malignant epithelial tumor is high-grade serous carcinoma, followed by endometrioid adenocarcinoma, clear cell carcinoma, and mucinous adenocarcinoma[19, 20]. Although most ovarian cancers arise as primary malignancies, approximately 10% are metastatic, often originating from breast, gastrointestinal, or gynecologic malignancies, such as endometrial carcinoma[21, 22].

Several studies have explored the relationship between a history of non-gynecologic cancers and the subsequent risk of developing ovarian cancer[23, 24]. A South Korean study found that individuals with colorectal cancer had a higher incidence of breast and gynecologic cancers than the general population, suggesting the role of genetic susceptibility, cancer-related treatments, environmental exposures, or hormonal factors in increasing risk[25]. Similarly, research indicates that women with a history of breast cancer have an elevated risk of developing second primary peritoneal, epithelial ovarian, and fallopian tube cancers compared to those without breast cancer[26].

While previous studies have assessed the incidence of secondary ovarian cancer[27, 28], limited research has systematically compared the clinicopathologic characteristics and survival outcomes between patients with primary versus second primary ovarian cancer. This study aims to examine the characteristics, treatment patterns, and survival outcomes among women with primary versus second primary ovarian malignancies. In addition, we seek to identify key factors associated with survival differences in these two groups, which may help guide future screening strategies, risk stratification, and personalized treatment approaches.

## Methods

### Data Source and Study Population

This retrospective cohort study utilized data from the Surveillance, Epidemiology, and End Results (SEER) 18 Registry[29], which collects cancer incidence and survival data from population-based registries representing approximately 36.7% of the U.S. population as of 2017[6] Data were extracted from the “Incidence - SEER 18 Registries, Nov 2021 Sub (2000–2021)” database. The study included women diagnosed with ovarian cancer as either a first primary or second primary malignancy, identified using International Classification of Diseases for Oncology, 3rd edition (ICD-O-3) codes. Eligible patients were required to have complete clinicopathological information and survival data, be aged 18 years or older, and have been actively followed up. Patients were excluded if their diagnosis was reported only through autopsy, death certificate, or clinical diagnosis without histologic confirmation or if they had two or more prior primary cancers.

### Primary Explanatory Variable

#### Defining Primary vs. Second Primary Ovarian Malignancies

The main explanatory variable was whether ovarian cancer was a primary or second primary malignancy. Primary ovarian malignancies were defined as the first recorded malignancy in a patient’s lifetime Second primary ovarian malignancies were defined as ovarian cancer diagnosed at least one year after a prior non-ovarian primary cancer, with cases that had the same site and histological subtype as the first malignancy excluded. This classification allowed for a comprehensive evaluation of potential differences in tumor characteristics, treatment response, and survival outcomes between these two groups.

### Outcome Variables

#### Overall Survival vs. Cancer-Specific Survival

The study examined overall survival (OS) and cancer-specific survival (CSS) as primary outcomes. OS was defined as the time from ovarian cancer diagnosis to death from any cause, while CSS was defined as the time from ovarian cancer diagnosis to death specifically due to ovarian cancer. For reporting consistency, the terms overall mortality (OM) and cancer-specific mortality (CSM) were used interchangeably with OS and CSS, respectively. This approach reflects the intrinsic relationship between these metrics, where a decrease in cancer-specific mortality corresponds to improved cancer-specific survival, and vice versa.

### FIGO Staging and Histologic Classification

Ovarian cancer staging was classified using the International Federation of Gynecology and Obstetrics (FIGO) system, which categorizes disease progression as follows: Stage I (tumor confined to the ovaries), Stage II (tumor spread to pelvic structures), Stage III (tumor spread to the peritoneum outside the pelvis and/or lymph nodes), and Stage IV (distant metastases beyond the peritoneal cavity). To enhance the accuracy of our analysis, histological subtypes were reclassified based on the WHO 2020 classification, distinguishing between high-grade serous carcinoma, low-grade serous carcinoma, mucinous adenocarcinoma, endometrioid carcinoma, clear cell adenocarcinoma, malignant Brenner tumor, and others. These adjustments ensured that invasive and non-invasive tumors were not analyzed together, thereby improving the scientific rigor of the study.

### Handling Missing FIGO Stage Data with Multiple Imputation

A total of 26.0% of patients had missing FIGO stage data. To maintain sample size and reduce potential bias, multiple imputation was applied. This method replaces missing values with statistically estimated values based on existing patient characteristics, thereby preventing the loss of valuable data and reducing selection bias. Multiple imputation strengthens statistical power and allows for more reliable and generalizable findings compared to complete-case analysis.

### Ethical Considerations and Data Access Statement

This study utilized data from the Surveillance, Epidemiology, and End Results (SEER) Program, a publicly available and fully de-identified dataset. As such, institutional review board (IRB) or ethics committee approval was not required. The data were accessed on May 10, 2024, for research purposes. At no point did the authors have access to personally identifiable information of participants.

### Statistical Analysis

Continuous variables were analyzed using means and standard deviations (SD) for normally distributed data, compared using the t-test, while non-normally distributed data were presented as medians with interquartile ranges and analyzed using the Mann-Whitney U test. Categorical variables were expressed as counts and percentages and compared using the chi-square test.

To examine the association between primary versus second primary ovarian cancer and survival outcomes, we performed Cox proportional hazards models for overall survival (OS) and Fine-Gray proportional hazards models for cancer-specific survival (CSS), accounting for competing risks. Results were presented as hazard ratios (HR) with 95% confidence intervals (CI), and statistical significance was set at P < 0.05.

To improve the accuracy of our estimates, we incorporated year-fixed effects to control for temporal changes in treatment and diagnostic advancements, state-fixed effects to account for regional variations in healthcare access and cancer treatment, and robust standard errors to adjust for heteroskedasticity and within-group correlation, ensuring more precise statistical inference. All analyses were conducted using STATA 16.

## Results

### Table 1: Patient Characteristics

A total of 27,308 women diagnosed with ovarian cancer between 2000 and 2021 were included in the study, with 23,132 (84.7%) having primary ovarian cancer and 4,176 (15.3%) having second primary ovarian cancer (Table 1). Women with second primary ovarian malignancies were older at diagnosis, with a greater proportion aged >64 years (44.6% vs. 35.6%, p<0.001) compared to those with primary ovarian cancer. Significant differences were observed in race/ethnicity distribution, with a higher proportion of White women among those with second primary ovarian cancer (71.3% vs. 66.0%, p<0.001). Hispanic (12.9% vs. 15.3%) and Asian/Pacific Islander women (10.1% vs. 12.5%) were less likely to have second primary ovarian malignancies (Table 1).

**Table 1:**
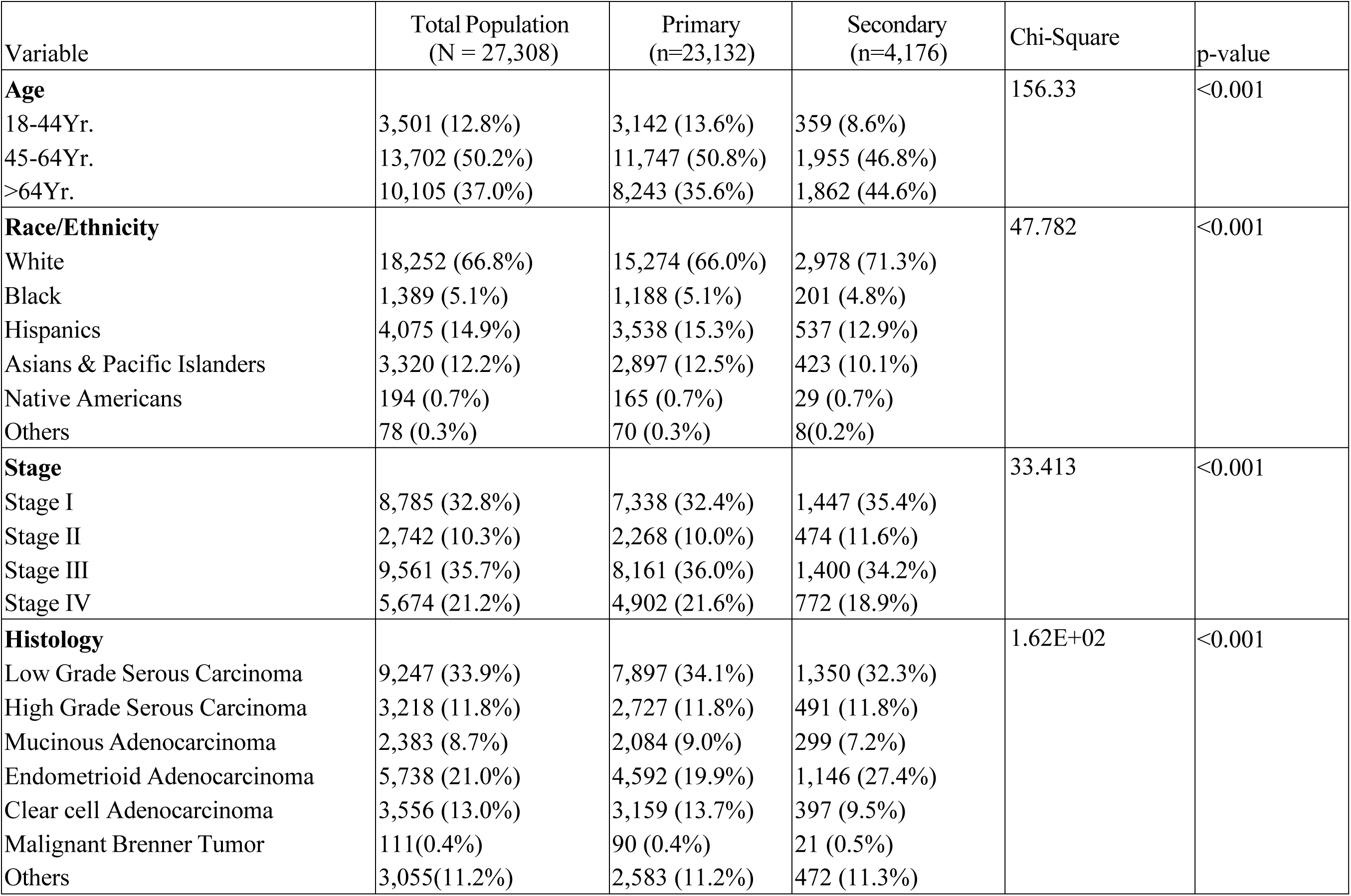
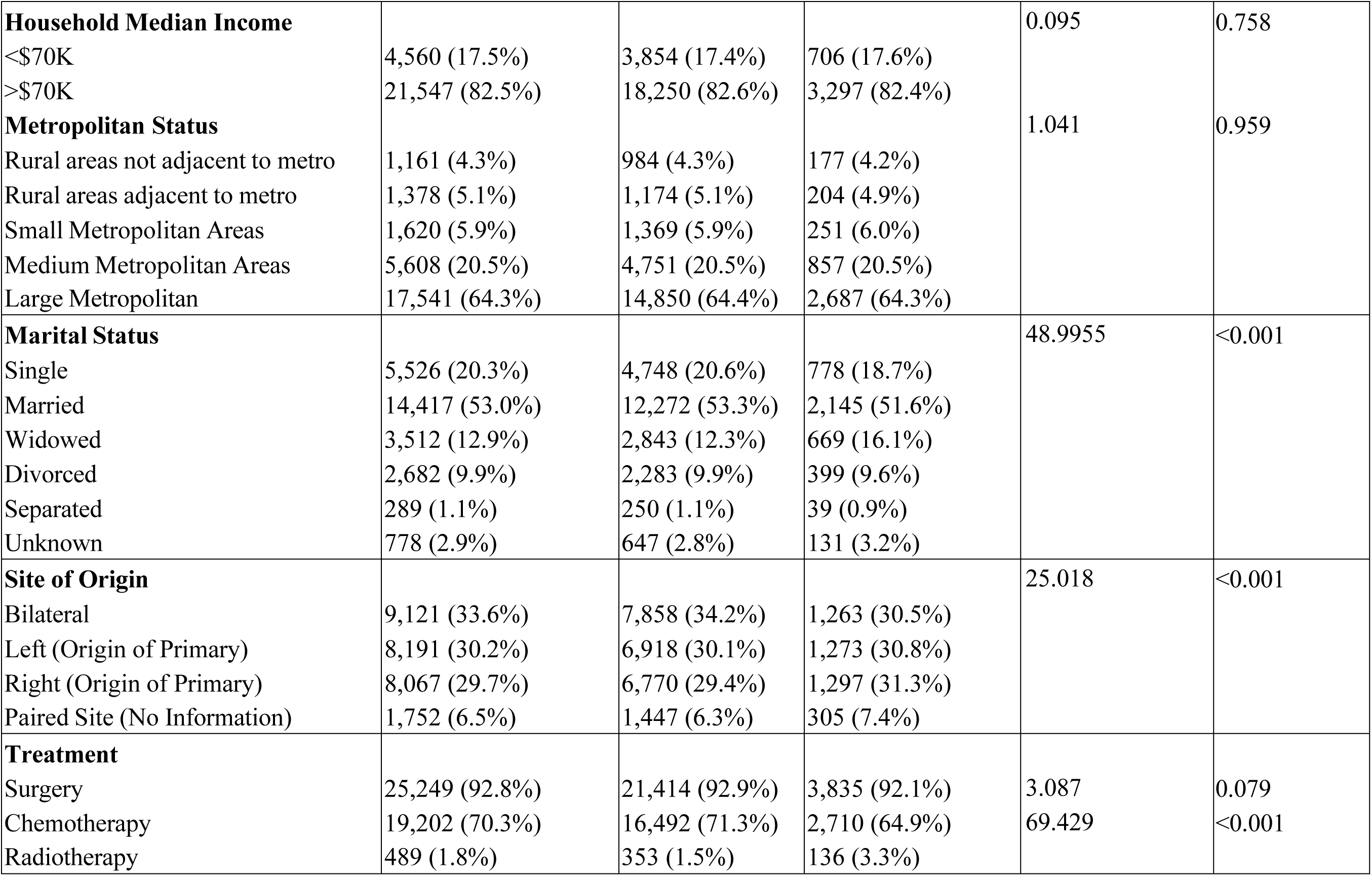
Baseline Characteristics of Women with Primary vs. Second Primary Ovarian Cancer: SEER Registry, 2000-2021.

#### Tumor Stage and Histologic Subtypes

The distribution of FIGO stage varied between the two groups (Table 1). Women with second primary ovarian cancer were more likely to be diagnosed at Stage I (35.4% vs. 32.4%) and Stage II (11.6% vs. 10.0%), while those with primary ovarian cancer were more likely to have Stage IV disease (21.6% vs. 18.9%, p<0.001). Histologic subtypes also differed between groups. Low-grade serous carcinoma was the most common histologic subtype in both groups (32.3% in second primary vs. 34.1% in primary ovarian cancer). Endometrioid adenocarcinoma was more frequently observed in the second primary group (27.4% vs. 19.9%), whereas clear cell adenocarcinoma was more prevalent in the primary ovarian cancer group (13.7% vs. 9.5%, p<0.001) (Table 1).

#### Socioeconomic and Geographic Factors

No significant differences were observed in household median income (Table 1), with 82.4% of second primary ovarian cancer patients and 82.6% of primary ovarian cancer patients having a household median income above $70,000 (p=0.758). Metropolitan status did not differ significantly between groups (p=0.959).

#### Marital Status and Tumor Site

Marital status varied between groups, with widowed women more likely to have second primary ovarian cancer (16.1% vs. 12.3%), while single (18.7% vs. 20.6%) and married women (51.6% vs. 53.3%) were slightly more prevalent in the primary ovarian cancer group (Table 1).

Regarding tumor site, bilateral ovarian involvement was more common in primary ovarian cancer (34.2% vs. 30.5%), while the distribution of left- and right-sided ovarian tumors was similar between groups (Table 1).

#### Treatment Patterns

Surgical treatment rates were similar between groups (Table 1), with 92.9% of primary ovarian cancer patients and 92.1% of second primary ovarian cancer patients undergoing surgery (p=0.079). However, chemotherapy use was significantly higher among women with primary ovarian cancer (71.3% vs. 64.9%, p<0.001). In contrast, radiotherapy was more frequently used in second primary ovarian cancer (3.3% vs. 1.5%, p<0.001) (Table 1).

### Table 2: Cancer-Specific and Overall Survival Rates in Primary vs. Second Primary Ovarian Cancer

#### Unadjusted Cancer-Specific Survival

Patients with second primary ovarian cancer demonstrated significantly better cancer-specific survival (CSS) than those with primary ovarian cancer across all time points (Table 2). The 5-year CSS rate was 60.3% in the second primary group compared to 56.8% in the primary group (p<0.001). This survival advantage persisted at 10 years (48.2% vs. 44.5%, p<0.001), 15 years (44.9% vs. 40.8%, p<0.001), and 20 years (43.5% vs. 39.1%, p<0.001) (Table 2).

**Table 2:** Unadjusted Cancer-Specific and Overall Survival Rates in Primary vs. Second Primary Ovarian Cancer.

| **Cancer-Specific Survival** | Total Population | Primary | Second Primary | p-value |
| --- | --- | --- | --- | --- |
| 5-Yr Survival | 57.3% | 56.8% | 60.3% | <0.001 |
| 10-Yr Survival | 45.1% | 44.5% | 48.2% | <0.001 |
| 15-Yr Survival | 41.4% | 40.8% | 44.9% | <0.001 |
| 20-Yr Survival | 39.7% | 39.1% | 43.5% | <0.001 |
| **Overall Survival** |  |  |  |  |
| 5-Yr Survival | 53.4% | 53.5% | 53.0% | 0.071 |
| 10-Yr Survival | 38.9% | 39.2% | 37.4% | <0.001 |
| 15-Yr Survival | 32.2% | 32.6% | 29.8% | <0.001 |
| 20-Yr Survival | 27.0% | 27.5% | 24.4% | <0.001 |

#### Unadjusted Overall Survival

In contrast to cancer-specific survival, overall survival (OS) was comparable between the two groups at 5 years (53.0% vs. 53.5%, p=0.071) (Table 2). However, the long-term OS trends showed slightly lower survival in the second primary ovarian cancer group. By 10 years, OS was 37.4% in the second primary group compared to 39.2% in the primary group (p<0.001). The survival gap widened further at 15 years (29.8% vs. 32.6%, p<0.001) and 20 years (24.4% vs. 27.5%, p<0.001) (Table 2).

### Table 3: Factors Associated with Cancer-Specific Survival

#### Primary vs. Second Primary Ovarian Cancer

Women with second primary ovarian cancer had a significantly lower hazard of cancer-specific mortality compared to those with primary ovarian cancer (HR: 0.928, 95% CI: 0.876-0.982, p=0.010), indicating a survival advantage among those with a prior malignancy (Table 3).

**Table 3:** Factors associated with Cancer-Specific Survival among Women with Ovarian Cancers: SEER Registry 2000-2021.

| Cancer-Specific Survival | Haz. Ratio | Std.Err | t | p-value | 95%CI |  |
| --- | --- | --- | --- | --- | --- | --- |
| Second Primary | 0.928 | 0.027 | -2.570 | 0.010 | 0.876 | 0.982 |
| **Age** |  |  |  |  |  |  |
| 18-44Yr. | Reference |  |  |  |  |  |
| 45/64Yr. | 1.356 | 0.050 | 8.210 | 0.000 | 1.261 | 1.458 |
| >64Yr. | 1.904 | 0.075 | 16.300 | 0.000 | 1.762 | 2.057 |
| **Race/Ethnicity** |  |  |  |  |  |  |
| White | Reference |  |  |  |  |  |
| Black | 1.328 | 0.059 | 6.370 | 0.000 | 1.217 | 1.449 |
| Hispanics | 0.993 | 0.030 | -0.210 | 0.831 | 0.936 | 1.055 |
| NHAPI | 0.900 | 0.035 | -2.720 | 0.007 | 0.834 | 0.971 |
| Native Americans | 1.330 | 0.144 | 2.630 | 0.009 | 1.075 | 1.644 |
| Unknown | 0.700 | 0.222 | -1.130 | 0.260 | 0.376 | 1.302 |
| **Histology** |  |  |  |  |  |  |
| Low Grade Serous Carcinoma | Reference |  |  |  |  |  |
| High Grade Serous Carcinoma | 0.967 | 0.032 | -1.010 | 0.314 | 0.907 | 1.032 |
| Mucinous Adenocarcinoma | 1.267 | 0.072 | 4.170 | 0.000 | 1.133 | 1.416 |
| Endometrioid Adenocarcinoma | 0.627 | 0.024 | -12.060 | 0.000 | 0.581 | 0.676 |
| Clear Cell Adenocarcinoma | 1.126 | 0.049 | 2.710 | 0.007 | 1.033 | 1.227 |
| Malignant Brenner Tumor | 0.914 | 0.214 | -0.380 | 0.703 | 0.577 | 1.449 |
| Others | 1.204 | 0.041 | 5.430 | 0.000 | 1.126 | 1.288 |
| **Site** |  |  |  |  |  |  |
| Bilateral | Reference |  |  |  |  |  |
| Left (Origin of Primary) | 0.714 | 0.021 | -11.510 | 0.000 | 0.674 | 0.756 |
| Right (Origin of Primary) | 0.729 | 0.021 | -11.020 | 0.000 | 0.689 | 0.771 |
| Paired Site (no Information) | 0.989 | 0.047 | -0.230 | 0.818 | 0.902 | 1.085 |
| **Stage** |  |  |  |  |  |  |
| Stage I | Reference |  |  |  |  |  |
| Stage II | 1.943 | 0.117 | 11.070 | 0.000 | 1.727 | 2.185 |
| Stage III | 3.891 | 0.195 | 27.090 | 0.000 | 3.526 | 4.294 |
| Stage IV | 4.625 | 0.260 | 27.290 | 0.000 | 4.142 | 5.163 |
| **Household Median Income** |  |  |  |  |  |  |
| <\$70k | 0.936 | 0.037 | -1.690 | 0.092 | 0.866 | 1.011 |
| **Married** |  |  |  |  |  |  |
| Single | Reference |  |  |  |  |  |
| Married | 0.912 | 0.026 | -3.280 | 0.001 | 0.863 | 0.963 |
| Widowed | 1.126 | 0.043 | 3.110 | 0.002 | 1.045 | 1.213 |
| Divorced | 1.051 | 0.040 | 1.320 | 0.185 | 0.976 | 1.132 |
| Separated | 1.066 | 0.114 | 0.590 | 0.554 | 0.863 | 1.315 |
| Unknown | 0.931 | 0.057 | -1.170 | 0.243 | 0.825 | 1.050 |
| **Metropolitan Status** |  |  |  |  |  |  |
| Rural not adjacent to metro | Reference |  |  |  |  |  |
| Rural adjacent to metro | 1.029 | 0.067 | 0.430 | 0.664 | 0.905 | 1.170 |
| Small Metropolitan | 0.941 | 0.062 | -0.920 | 0.357 | 0.827 | 1.071 |
| Medium Metropolitan | 0.970 | 0.058 | -0.500 | 0.616 | 0.862 | 1.092 |
| Large Metropolitan | 0.937 | 0.061 | -1.010 | 0.311 | 0.825 | 1.063 |
| **Treatment** |  |  |  |  |  |  |
| Chemotherapy | 1.029 | 0.030 | 0.980 | 0.328 | 0.972 | 1.090 |
| Surgery | 0.369 | 0.018 | -20.250 | 0.000 | 0.335 | 0.406 |
| Radiotherapy | 1.024 | 0.079 | 0.310 | 0.760 | 0.880 | 1.191 |

*Model adjusted for state-fixed effects and year-fixed effects.*

#### Age and Race/Ethnicity

Age at diagnosis was a strong predictor of cancer-specific survival (Table 3). Compared to younger women (18-44 years), those aged 45-64 years had a 35.6% higher risk of cancer-related mortality (HR: 1.356, 95% CI: 1.261-1.458, p<0.001), while those aged >64 years had a 90.4% higher risk (HR: 1.904, 95% CI: 1.762-2.057, p<0.001).

Race/ethnicity was also associated with CSS. Compared to White women, Black women had a 32.8% higher risk of cancer-specific mortality (HR: 1.328, 95% CI: 1.217-1.449, p<0.001), while Native American women also exhibited an increased risk (HR: 1.330, 95% CI: 1.075-1.644, p=0.009). In contrast, Asian/Pacific Islander women (NHAPI) had improved CSS (HR: 0.900, 95% CI: 0.834-0.971, p=0.007) (Table 3).

#### Histologic Subtype and Tumor Site

Histologic subtype significantly influenced CSS. Compared to low-grade serous carcinoma (Table 3), mucinous adenocarcinoma (HR: 1.267, 95% CI: 1.133-1.416, p<0.001) and clear cell adenocarcinoma (HR: 1.126, 95% CI: 1.033-1.227, p=0.007) were associated with poorer survival. Conversely, endometrioid adenocarcinoma was associated with better survival (HR: 0.627, 95% CI: 0.581-0.676, p<0.001).

Tumor site of origin also impacted survival. Compared to bilateral ovarian cancer, left-sided tumors (HR: 0.714, 95% CI: 0.674-0.756, p<0.001) and right-sided tumors (HR: 0.729, 95% CI: 0.689-0.771, p<0.001) were associated with improved survival (Table 3).

#### FIGO Stage and Marital Status

Cancer stage at diagnosis was a strong predictor of CSS. Compared to Stage I (Table 3), the risk of cancer-specific mortality was 94.3% higher in Stage II (HR: 1.943, 95% CI: 1.727-2.185, p<0.001), nearly four times higher in Stage III (HR: 3.891, 95% CI: 3.526-4.294, p<0.001), and over four and a half times higher in Stage IV (HR: 4.625, 95% CI: 4.142-5.163, p<0.001).

Marital status was also associated with survival. Compared to single women, married women had improved CSS (HR: 0.912, 95% CI: 0.863-0.963, p=0.001), whereas widowed women had a higher risk of cancer-specific mortality (HR: 1.126, 95% CI: 1.045-1.213, p=0.002) (Table 3).

#### Treatment Modalities and Metropolitan Status

Among treatment modalities, surgery was the most significant predictor of improved CSS, with a 63.1% reduction in cancer-specific mortality (HR: 0.369, 95% CI: 0.335-0.406, p<0.001). In contrast, chemotherapy (HR: 1.029, p=0.328) and radiotherapy (HR: 1.024, p=0.760) were not significantly associated with CSS (Table 3).

Metropolitan status did not significantly impact survival, with no differences observed between rural and metropolitan residents (p>0.05) (Table 3).

### Table 4: Factors Associated with Overall Survival

#### Primary vs. Second Primary Ovarian Cancer

Women with second primary ovarian cancer had a 5.7% higher risk of overall mortality compared to those with primary ovarian cancer (HR: 1.057, 95% CI: 1.005-1.113, p=0.031), suggesting a slight survival disadvantage for those with second primary malignancies (Table 4).

**Table 4:** Factors associated with Overall Survival among Women with Ovarian Cancers: SEER Registry 2000-2021.

| Overall Survival | Haz. Ratio | Std.Err | t | p-value | 95%CI |  |
| --- | --- | --- | --- | --- | --- | --- |
| Second Primary | 1.057 | 0.027 | 2.160 | 0.031 | 1.005 | 1.113 |
| **Age** |  |  |  |  |  |  |
| 18-44Yr. | Reference |  |  |  |  |  |
| 45/64Yr. | 1.471 | 0.053 | 10.680 | 0.000 | 1.370 | 1.579 |
| >64Yr. | 2.394 | 0.091 | 23.040 | 0.000 | 2.223 | 2.578 |
| **Race/Ethnicity** |  |  |  |  |  |  |
| White | Reference |  |  |  |  |  |
| Black | 1.301 | 0.055 | 6.260 | 0.000 | 1.198 | 1.413 |
| Hispanics | 1.015 | 0.029 | 0.500 | 0.615 | 0.959 | 1.073 |
| NHAPI | 0.890 | 0.032 | -3.210 | 0.001 | 0.828 | 0.955 |
| Native Americans | 1.275 | 0.131 | 2.370 | 0.018 | 1.043 | 1.559 |
| Unknown | 0.702 | 0.201 | -1.240 | 0.217 | 0.401 | 1.231 |
| **Histology** |  |  |  |  |  |  |
| Low Grade Serous Carcinoma | Reference |  |  |  |  |  |
| High Grade Serous Carcinoma | 0.967 | 0.030 | -1.080 | 0.281 | 0.910 | 1.028 |
| Mucinous Adenocarcinoma | 1.323 | 0.068 | 5.480 | 0.000 | 1.197 | 1.463 |
| Endometrioid Adenocarcinoma | 0.712 | 0.023 | -10.300 | 0.000 | 0.668 | 0.760 |
| Clear Cell Adenocarcinoma | 1.104 | 0.044 | 2.470 | 0.013 | 1.021 | 1.194 |
| Malignant Brenner Tumor | 1.128 | 0.203 | 0.670 | 0.504 | 0.792 | 1.606 |
| Others | 1.189 | 0.038 | 5.360 | 0.000 | 1.116 | 1.267 |
| **Site** |  |  |  |  |  |  |
| Bilateral | Reference |  |  |  |  |  |
| Left (Origin of Primary) | 0.746 | 0.020 | -11.030 | 0.000 | 0.708 | 0.786 |
| Right (Origin of Primary) | 0.762 | 0.020 | -10.450 | 0.000 | 0.724 | 0.802 |
| Paired Site (no Information) | 0.999 | 0.046 | -0.030 | 0.974 | 0.913 | 1.092 |
| **Stage** |  |  |  |  |  |  |
| Stage I | Reference |  |  |  |  |  |
| Stage II | 1.593 | 0.076 | 9.760 | 0.000 | 1.451 | 1.750 |
| Stage III | 2.930 | 0.118 | 26.640 | 0.000 | 2.707 | 3.172 |
| Stage IV | 3.450 | 0.161 | 26.470 | 0.000 | 3.147 | 3.782 |
| Household Median Income<br><\$70K | 0.911 | 0.034 | -2.490 | 0.013 | 0.846 | 0.980 |
| **Married** |  |  |  |  |  |  |
| Single | Reference |  |  |  |  |  |
| Married | 0.897 | 0.024 | -4.110 | 0.000 | 0.852 | 0.945 |
| Widowed | 1.160 | 0.040 | 4.300 | 0.000 | 1.084 | 1.241 |
| Divorced | 1.086 | 0.038 | 2.330 | 0.020 | 1.013 | 1.164 |
| Separated | 1.069 | 0.109 | 0.650 | 0.513 | 0.876 | 1.304 |
| Unknown | 0.932 | 0.054 | -1.200 | 0.230 | 0.832 | 1.045 |
| **Metropolitan Status** |  |  |  |  |  |  |
| Rural not adjacent to metro | Reference |  |  |  |  |  |
| Rural adjacent to metro | 1.010 | 0.060 | 0.160 | 0.871 | 0.898 | 1.135 |
| Small Metropolitan | 0.931 | 0.057 | -1.170 | 0.243 | 0.825 | 1.050 |
| Medium Metropolitan | 0.960 | 0.053 | -0.750 | 0.455 | 0.862 | 1.069 |
| Large Metropolitan | 0.929 | 0.055 | -1.240 | 0.216 | 0.827 | 1.044 |
| **Treatment** |  |  |  |  |  |  |
| Chemotherapy | 0.951 | 0.024 | -1.980 | 0.048 | 0.904 | 1.000 |
| Surgery | 0.378 | 0.018 | -20.140 | 0.000 | 0.344 | 0.416 |
| Radiotherapy | 1.020 | 0.073 | 0.280 | 0.781 | 0.886 | 1.174 |

*Model adjusted for state-fixed effects and year-fixed effects.*

#### Age and Race/Ethnicity

Older age at diagnosis was a strong predictor of worse overall survival (Table 4). Compared to women aged 18-44 years, those aged 45-64 years had a 47.1% higher risk of mortality (HR: 1.471, 95% CI: 1.370-1.579, p<0.001), while those older than 64 years had a more than twofold increased risk (HR: 2.394, 95% CI: 2.223-2.578, p<0.001).

Race/ethnicity was also significantly associated with OS. Black women had a 30.1% increased risk of overall mortality (HR: 1.301, 95% CI: 1.198-1.413, p<0.001), while Native American women also exhibited a higher risk (HR: 1.275, 95% CI: 1.043-1.559, p=0.018). In contrast, non-Hispanic Asian/Pacific Islander (NHAPI) women had improved OS (HR: 0.890, 95% CI: 0.828-0.955, p=0.001) (Table 4).

#### Histologic Subtype and Tumor Site

Histologic subtype had a significant impact on survival (Table 4). Compared to low-grade serous carcinoma, mucinous adenocarcinoma (HR: 1.323, 95% CI: 1.197-1.463, p<0.001), clear cell adenocarcinoma (HR: 1.104, 95% CI: 1.021-1.194, p=0.013), and other histologic subtypes (HR: 1.189, 95% CI: 1.116-1.267, p<0.001) were associated with worse OS. Conversely, endometrioid adenocarcinoma was associated with better survival (HR: 0.712, 95% CI: 0.668-0.760, p<0.001).

Regarding tumor site of origin, compared to bilateral ovarian involvement, left-sided tumors (HR: 0.746, 95% CI: 0.708-0.786, p<0.001) and right-sided tumors (HR: 0.762, 95% CI: 0.724-0.802, p<0.001) were associated with improved overall survival (Table 4).

#### FIGO Stage and Marital Status

Advanced disease stage was strongly associated with worse OS (Table 4). Compared to Stage I, patients with Stage II disease had a 59.3% increased risk of mortality (HR: 1.593, 95% CI: 1.451-1.750, p<0.001), while those with Stage III (HR: 2.930, 95% CI: 2.707-3.172, p<0.001) and Stage IV (HR: 3.450, 95% CI: 3.147-3.782, p<0.001) had significantly worse survival outcomes.

Marital status also influenced survival. Married women had better OS (HR: 0.897, 95% CI: 0.852-0.945, p<0.001) compared to single women, whereas widowed women had a significantly higher risk of mortality (HR: 1.160, 95% CI: 1.084-1.241, p<0.001) (Table 4).

#### Treatment Modalities and Metropolitan Status

Among treatment modalities, surgery was the strongest predictor of improved OS (Table 4), reducing mortality risk by 62.2% (HR: 0.378, 95% CI: 0.344-0.416, p<0.001). Chemotherapy was marginally associated with improved survival (HR: 0.951, 95% CI: 0.904-1.000, p=0.048), while radiotherapy did not significantly impact OS (HR: 1.020, 95% CI: 0.886-1.174, p=0.781). Metropolitan status was not significantly associated with OS, with no substantial differences observed between rural and urban populations (p>0.05) (Table 4).

#### Trends in Low-Grade and High-Grade Serous Carcinoma

The incidence of low-grade serous carcinoma (LGSC) and high-grade serous carcinoma (HGSC), the two most common epithelial ovarian cancer subtypes, demonstrated opposite trends over time (Figure 1). In 2000, LGSC accounted for 52.64% of serous carcinomas, whereas HGSC represented only 7.9%. Over the next two decades, LGSC incidence steadily declined, reaching 7.43% by 2021. In contrast, HGSC incidence remained relatively low until 2016, after which it increased sharply, surpassing LGSC by 2018 (22.42%) and reaching its highest recorded incidence of 33.31% in 2021(Figure 1).

**Figure 1.** Trends in the incidence of low-grade serous carcinoma (LGSC) and high-grade serous carcinoma (HGSC) from 2000 to 2021, showing a decline in LGSC and a sharp increase in HGSC after 2015.

#### Cancer-Specific Survival in Primary vs. Second Primary Ovarian Malignancies

The Kaplan-Meier survival analysis (Figure 2) demonstrated a significant difference in cancer-specific survival between primary ovarian cancers and second primary ovarian malignancies (log-rank test, p < 0.001). Patients with second primary ovarian malignancies exhibited higher survival probabilities over time compared to those with primary ovarian cancer. At 5 years, the survival probability remained higher in the second primary group, and this trend persisted throughout the 20-year follow-up period. The number at risk progressively declined in both groups, but the survival advantage for second primary malignancies remained evident (Figure 2).

**Figure 2.** A Kaplan-Meier survival curve comparing cancer-specific survival between Primary ovarian cancers and Second Primary ovarian malignancies using data from the SEER registry (2000-2021). The survival probability is plotted over time (years). The log-rank test (x - 15.19, p < 0.001) indicates a statistically significant difference in survival between the two groups, with Second Primary ovarian malignancies demonstrating better survival outcomes. The number at risk table below shows the number of patients at risk at different time points.

#### Overall Survival in Primary vs. Second Primary Ovarian Cancers

The Kaplan-Meier survival analysis (Figure 3) revealed a statistically significant difference in overall survival between primary ovarian cancers and second primary ovarian malignancies (log-rank test, p = 0.018). While the survival curves remained closely aligned, patients with primary ovarian cancer demonstrated a slightly higher overall survival probability over time compared to those with second primary ovarian malignancies. The survival difference between the two groups was less pronounced than in cancer-specific survival, suggesting that non-cancer-related factors, such as comorbidities or treatment history, may contribute to higher long-term mortality in second primary malignancies. The number at risk table indicates a progressive decline in patient counts across both groups, with a more gradual survival reduction in primary ovarian cancers (Figure 3).

**Figure 3.** A Kaplan-Meier survival curve comparing overall survival between Primary ovarian cancers and Second Primary ovarian malignancies using data from the SEER registry (2000-2021). The survival probability is plotted over time (years). The log-rank test (° - X.XX, p < 0.018) suggests a statistically significant difference in overall survival between the two groups, though the magnitude of difference is smaller than in cancer-specific survival. The number at risk table below indicates the number of patients at risk at various time points.

## Discussion

This study provides important insights into the survival differences between primary ovarian cancer and second primary ovarian cancer, as well as the evolving incidence of serous carcinoma subtypes. Women with second primary ovarian cancer demonstrated higher CSS than those with primary ovarian malignancies. However, OS was lower among women with second primary ovarian cancer, likely due to the cumulative burden of multiple malignancies over a lifetime. In addition, our findings reveal a shifting epidemiology in serous carcinoma, where low-grade serous carcinoma (LGSC) was historically the predominant subtype but was overtaken by high-grade serous carcinoma (HGSC) after 2018.

Consistent with previous literature, older age, advanced disease stage, and Black race were associated with higher cancer-specific and overall mortality, reflecting disparities in access to care, biological differences, and comorbid disease burden[30–35]. Marital status conferred a protective effect, with married women exhibiting improved survival compared to unmarried women[36–38]. In addition, among histologic subtypes, mucinous adenocarcinoma was associated with the worst survival outcomes[39, 40].

Our findings align with previous studies suggesting that second primary ovarian malignancies may have distinct biological behaviors and management strategies, leading to improved cancer-specific survival[41–43]. Several mechanisms may explain the observed survival differences. The improved CSS in second primary ovarian malignancies likely results from heightened clinical vigilance, routine surveillance, and early detection[44, 45]. Women with a prior cancer diagnosis often undergo regular medical follow-ups, leading to earlier-stage diagnosis and timely intervention, which may contribute to better ovarian cancer-specific outcomes[44]. However, their worse OS may stem from cumulative exposure to chemotherapy, radiotherapy, or surgery, leading to organ dysfunction, treatment resistance, or secondary complications[41, 46].

The shift in serous carcinoma epidemiology is noteworthy. Our study reveals a significant decline in the incidence of low-grade serous carcinoma over time, while high-grade serous carcinoma has become more prevalent, surpassing LGSC by 2018. This trend may be attributed to advancements in diagnostic techniques, improved molecular profiling, and a better understanding of HGSC as a distinct entity with unique therapeutic implications[47]. Given that HGSC is associated with higher genomic instability, BRCA mutations, and aggressive clinical behavior, the observed increase in its incidence underscores the importance of enhanced screening, tailored treatment strategies, and risk-reduction approaches for high-risk individuals[48–50].

### Implications for Population Health and Policy

These findings have significant public health and clinical implications. The improved CSS but worse OS in second primary ovarian cancer patients suggests a need for integrated survivorship programs that not only focus on cancer recurrence surveillance but also address long-term comorbidities, treatment-related toxicities, and quality-of-life concerns. In addition, the rising burden of HGSC calls for enhanced genetic testing, early detection programs, and improved therapeutic strategies to mitigate its aggressive course. Given the disparities in survival outcomes by race and geographic location, targeted interventions are necessary to reduce healthcare access inequalities. Policies should focus on expanding access to genetic counseling, increasing participation in clinical trials, and implementing community-based initiatives to support early diagnosis and treatment adherence, particularly among minority populations.

### Limitations and Future Directions

This study utilizes data from the SEER registry, which provides a robust, population-based dataset. However, several limitations must be acknowledged:

#### Lack of First Primary Tumor and Germline Mutation Data

The SEER database does not provide information on the first primary malignancy in cases of second primary ovarian cancers, limiting our ability to assess the impact of prior cancer screening and treatment on subsequent ovarian malignancy survival. Additionally, germline pathogenic mutations (e.g., Li-Fraumeni syndrome, Peutz-Jeghers syndrome, and hereditary breast-ovarian cancer syndrome) were not available, despite their potential influence on survival outcomes.

#### Timeframe and the Impact of PARP Inhibitors (PARPi)

Our study period now extends from 2000 to 2021, addressing prior concerns about capturing long-term survival trends. However, while we included year fixed effects to account for treatment advancements, our dataset may not fully capture the long-term impact of PARPi, which became widely used after 2019 and has significantly improved progression-free survival in BRCA1/2 mutation carriers.

#### Use of FIGO Staging and Histological Reclassification

In response to reviewer concerns, we have reclassified tumor grade using the latest FIGO staging system and adopted the WHO 2020 histological classification for ovarian cancer. While this standardization improves study validity, it led to a significant reduction in sample size (from 97,288 to 27,308 cases), potentially impacting the generalizability of our findings.

#### Lack of Surgical and Targeted Therapy Data

SEER does not provide details on cytoreductive surgery outcomes, including R0 resection status, which is a critical determinant of prognosis in ovarian cancer. Additionally, chemotherapy regimens and targeted therapies, including PARPi use, were not available, preventing a detailed assessment of their influence on survival disparities.

#### Comorbidities and Performance Status

Our analysis did not include data on comorbidities, functional status, or performance scores, which could influence treatment decisions and overall survival outcomes. While we adjusted for baseline characteristics, disease stage, and treatment modalities, residual confounding due to unmeasured health status factors may still be present.

Despite these limitations, our study provides important insights into survival differences between primary and second primary ovarian cancers, utilizing a large, population-based dataset. Future research should focus on integrating molecular and treatment-specific data to further elucidate survival disparities in ovarian cancer.

## Conclusion

Our study provides critical insights into survival differences between primary and second primary ovarian cancers, revealing higher CSS but worse OS in second primary ovarian malignancies. We also document a notable shift in serous carcinoma epidemiology, with HGSC surpassing LGSC as the predominant subtype after 2018. These findings highlight the importance of early detection, improved treatment strategies, and equitable healthcare policies to optimize outcomes for ovarian cancer patients. Future research should explore the underlying molecular mechanisms and identify interventions to further improve survival, particularly in high-risk populations.

## Data Availability

All relevant data are within the manuscript and its Supporting Information files.

## Supporting Information

**S1 Fig. Trends in low grade and high-grade serous carcinoma incidence (2000-2021).**

**S2 Fig. Cancer specific survival in primary vs secondary ovarian malignancies.**

**S3 Fig. Overall survival in primary vs secondary ovarian cancer.**

